# How to quit confinement? French scenarios face to COVID-19

**DOI:** 10.1101/2020.04.02.20051342

**Authors:** Augeraud-Véron Emmanuelle

## Abstract

A mathematical model is developed to study the spread of the COVID-19 epidemics in France. Data from French Public Agency of Health are considered to calibrate the model. The spread of the epidemics strongly depends on confinement measures. The aim of the paper is to predict the evolution of the epidemics under various scenarios that could be taken to quit confinement. The spread of the disease and its re-emergence strongly depends on these scenarios.

## 1 Introduction

In December 2019, the World Health Organization warned of several cases of pneumonia in Wuhan, China. On January 7th, 2020, China confirmed that it was a new virus of the corona virus family called COVID-19. On March 11th, the World Health Organization officially declared a pandemic. In March 22nd, there were 81500 confirmed cases in China and 3267 deaths. 188 countries in the world were affected by this date. However, on March 18th, 2020, China declared that no new contaminations had been reported for the first time since the beginning of the epidemic. This stop of the spread of the epidemics has been made possible thanks to confinement. Indeed, at the end of January 2020, due to the extend of the spread of the virus on Chinese territory, China put in place drastic confinement measures.

In France, the corona virus pandemic begun on January 24th 2020, when the first three cases were recorded. As 100 individuals were already reported as having the disease and 2 deaths, stade 2 of ORSAN plan was activated of February the 29th. It’s aim is to slow down the spread of the epidemics. With 4500 reported cases and 91 deaths on March 14th, the epidemic situation moved to stage 3 and all places receiving non-essential public traffic have been closed. Since March the 17th, the population is confined to their homes, except for authorized reasons and movement is limited to a strict minimum.

In March the 25th, it is estimated that more than 100,000 fines have been issued for non-compliance with confinement (Mignon Colombet and Floreancig [8]). Since March the 17th, the arsenal against confinement violators is getting stronger. A fine of 135 euros has first been instituted for violation of travel bans. Two days later, five people who had been fined several times for non respect of confinement were taken into police custody for *endangering the lives of others*. The government has had to take more coercive measures in the face of non-compliance with confinement constraints. The correctional court had to impose a prison sentence on the basis of endangerment of life. (Article 223-1 of French Penal Code).

On the 27th of March, confinement initially scheduled to the end of March has been prolongated to April the 15th by Prime Minister Edouard Phillipe who also indicated that the situation will be re-examined according to new reported and death cases, while in the same time the Scientific Council, for its part, estimated the duration of the confinement should be *six weeks* long from the start of confinement.

The question can be asked to understand the future of the epidemics when the confinement will be released, as the number of new reported cases depends strongly on this confinement measures. The impact of quitting confinement scenarios on the spread of the disease will be studied.

## 1.1 Results

### 1.2 The model and data

We consider a SEIR model with differential infectivity. The spread of the disease across the population is considered in System 1.

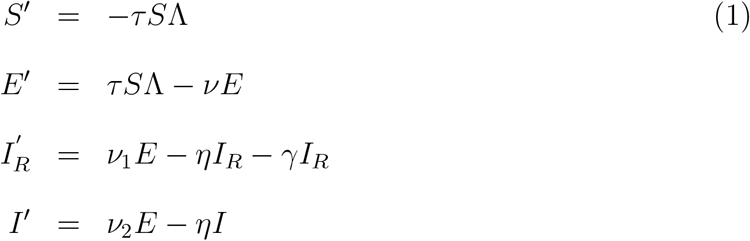

With

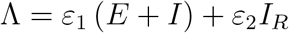

with initial data

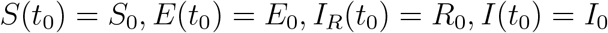

*S*(*t*) represents the number of individuals susceptible to be contaminated by the infection. At time *t > t*_0_, susceptible may be contaminated by asymptomatic infectious individuals *E*(*t*) (also called exposed). Individual stay exposed for a mean time duration *v*^−1^. Either they become lightly asymptomatic *I* at rate *v*_2_ either they present enough symptoms and are reported *I*_*R*_ at rate *v*_1_. Individuals stay *I*_*R*_ or *I* during a period *η*^−1^ in average. When confinement is imposed, contamination rate is reduced from a factor *ε*_1_ ∈ [0, 1]. This

parameter depends on health policy, but also on behavioral component (protections that individuals really apply). We would consider this parameter as a control parameter in order to study the spread of the epidemics for different values of *ε*. In subsection 1.4 we would consider that this parameter is time dependent in order to take into account health policy measures. In stage two of the ORSAN plan, this would model the change of contact habit between individuals and the adoption of the barrier gesture. In stage 3, it will model the confinement, which took time to be respected by individuals. *ε*_2_ ∈ [0, 1] is due to recommendation of isolation that is required by doctors in reported cases. It is assumed that reported cases may still be contaminant, for example while they are being cared for in the hospital.

The present modelisation is based on Liu et al. ([5],[6]) who model the spread of the epidemics in Wuhan and in China and on Magal and Webb ([7]) who calibrate the epidemics with early data on several countries among which France. However, our modelisation differs in two points from theirs. Firstly, they do not consider contamination by reported symptomatic cases. It appears that in France, many cases of contamination happens in hospital (in France, on March 22nd, the first case of a medical doctor has been announced). Secondly, we also consider death due to the disease.

We would use calibration obtained in Magal and Webb ([7]) to describe the epidemics before the application of confinement measures. We recalibrate the impact of confinement on the force of infection taking into account more recent data. Our aim in this paper is to focus on the impact of leaving-confinement policy and behavioral changes.

To describe the evolution of the epidemics, we consider the following parameters:

| Symbol | Interpretation | Value |
| --- | --- | --- |
| $t_0$ | Time at which the epidemics started | 0 |
| $t_1$ | Time at which the quarantine started | 11 [[9]] |
| $t_2$ | Time at which the quarantine ends | 49 |
| $S_0$ | Number of susceptible at time $t_0$ | $66.99.10^6$ |
| $E_0$ | Number of asymptomatic infectious at time $t_0$ | 4.2 [[7]] |
| $I_0$ | Number of unreported symptomatic infectious cases at time $t_0$ | 0.58 [[7]] |
| $I_{R,0}$ | Number of reported symptomatic infectious cases at time $t_0$ | 0 |
| $\tau$ | transmission rate | $6.78.10^{-9}$ [[7]] |
| $\gamma$ | Death due to the disease | fitted |
| $\nu^{-1}$ | Average duration of the asymptomatic infectious period | 7 |
| $f$ | Fraction of asymptomatic infectious who become reported symptomatic infectious | 0.6 |
| $\nu_1 = f\nu$ | Rate at which asymptomatic infectious become reported symptomatic infectious | |
| $\nu_2 = (1 - f)\nu$ | Rate at which asymptomatic infectious become unreported symptomatic infectious | |
| $\eta^{-1}$ | Average time symptomatic infectious have symptoms | 7 |
| $\varepsilon_1(t)$ | intensity of contacts among unreported infected individuals at time $t$ | fitted |
| $\varepsilon_2$ | intensity of contacts among reported infected individuals | 0.0001 |

### 1.3 Final size of the epidemics

To determine the size of the epidemics we would consider a new variable *Y* computed as a weighted sum of people involved in the transmission of the epidemics. This approach has been developed by Arino et al. [1] in a general framework applied to various cases and Feng [2] in a SEIR model where the exposed class does not propagate the virus. Our modeling computation is detailed in the Supplementary Material section.

Let us denote *S*_∞_ = lim_*t*→∞_ *S* (*t*) and *Y*_∞_ = lim_*t*→∞_ *Y* (*t*),. Then *Y*_∞_ = 0, and computations show^1^ that the final size of the epidemics is obtained by the relation

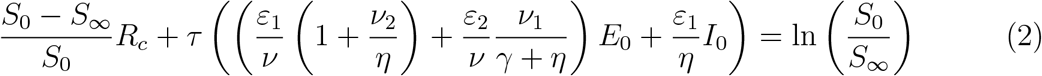

where *ℛ*_*C*_ is the controlled reproduction number, which is computed from System (1) as

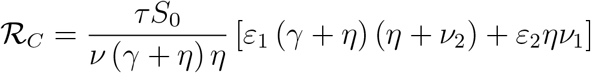

Equation (2) can also be expressed in terms of individuals who have been infected during the whole duration of the epidemics. Denoting *C*_∞_ those infected individuals, then *C*_∞_ which is formally defined as *C*_∞_ = *S*_0_ − *S*_∞_ and satisfies

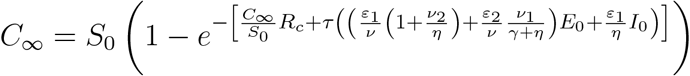

The final size of the epidemics strongly depends on the parameters of the model, and in particular on *ε*_1_.

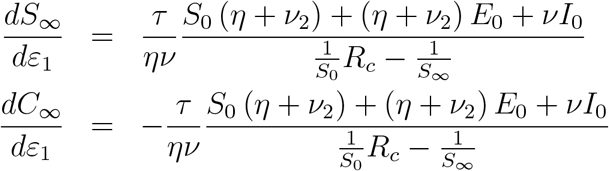

We can also compute the relative variation of the final number of non contaminated (or contaminated) according to the relative variation of *ε*_1_, which is called elasticity.

Elasticity 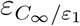, which is defined as

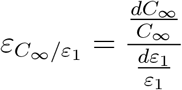

indicates that if *ε*_1_ increases of 1%, the number of whole individuals contaminated by the virus is 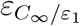. Elasticity 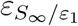 is defined in the same manner by

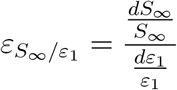

According to Equation 2 theses elasticity can be explicitly computed as

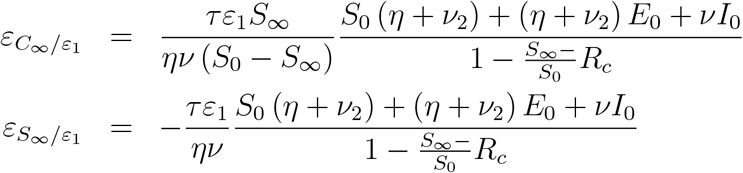

Thus the impact of *ϵ*_1_ on the final size of the epidemics can be studied. Non surprisingly in can be seen Figure 1 (a) the more contact among individuals the smallest the reservoir of non contaminated individuals at the end of the epidemics. As 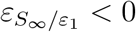, Figure 1 (b) indicates the same result. However, Figure 1 (b) also shows that elasticity 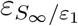 is not monotonous function of *ε*_1_. For *ε*_1_ closed to 0.28, a small variation of *ε*_1_ in percentage may have less impact on the final size of the epidemics than for greater or lower values of *ε*_1_. Figure 1 (c) shows elasticity 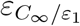 for different values of *ε*_1_. As 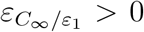, non surprisingly, the number of contaminated individuals during the course of the epidemics is an increasing function of contact *ε*_1_ among individuals. However, the smaller *ε*_1_ the stronger this impact.

**Figure 1:**
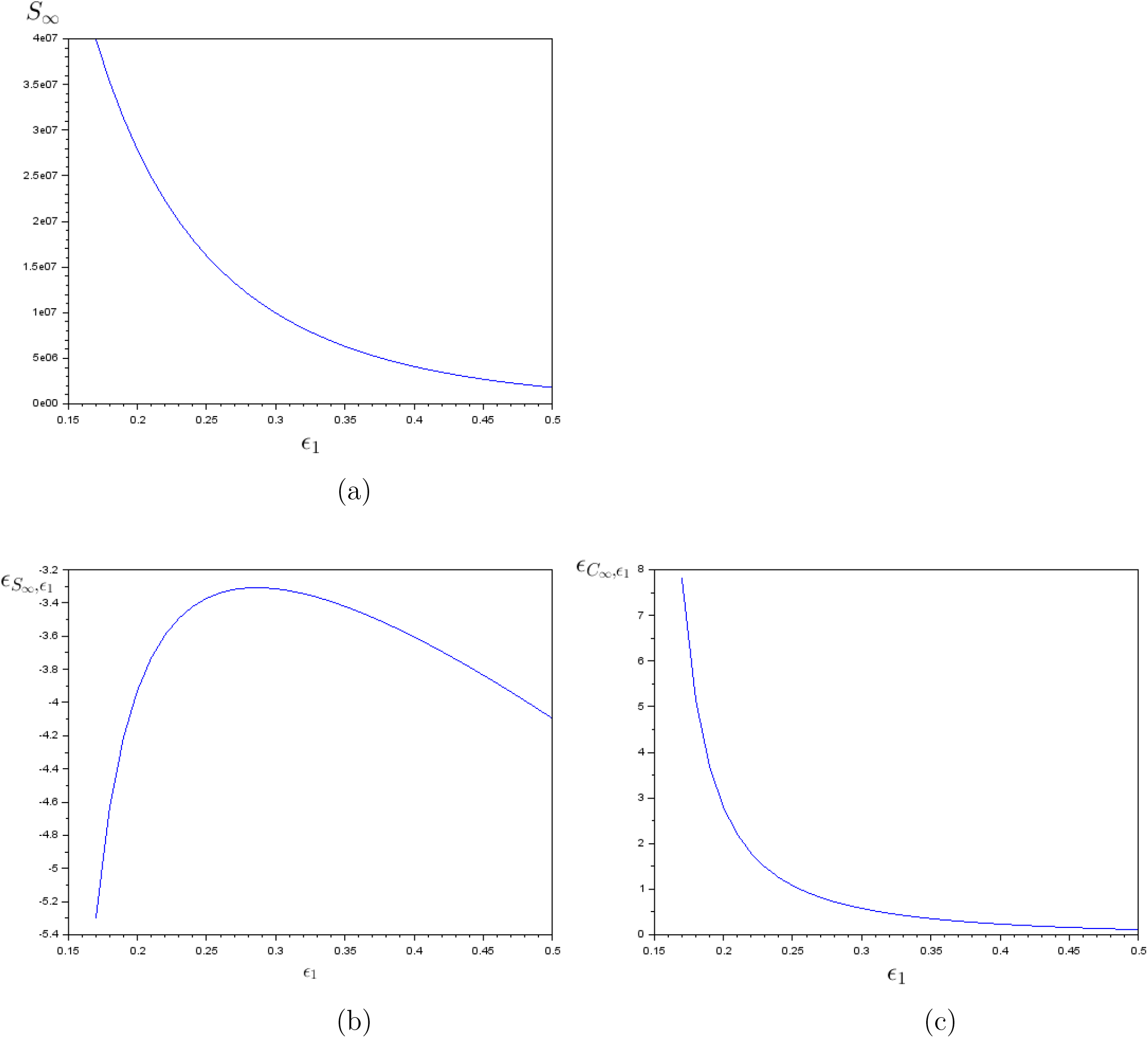
Impact of *ϵ*_1_, intensity of the contact among individuals, on the reservoir of non infected individuals at the end of the epidemics (a), on the elasticity of this reservoir relative to a variation of *∈*_1_ (b), and on elasticity of infected individuals at the end of the epidemics (c). Computed for *τ* = 6.78 ∗ 10 (− 9)

### 1.4 Numerical experiments

To take into account the slow awareness of individuals about the importance of confinement, we would assume that

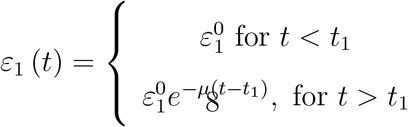

This function has been used in Liu et al. ([6],[5]). *µ* has been fitted to correspond to data of French Public Agency of Health. It was found that *µ* = 0.035.

Different scenario of disconfinement are now presented from time *t*_2_. On the difference to Ferguson et al. [4], we only consider one time of application of confinement and confinement-leaving.

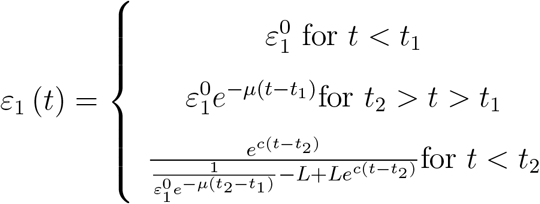

A fast disconfinement scenario is first considered corresponding to 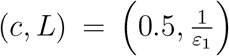. slow disconfinement scenario is then considered with 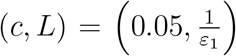. In this two scenarios, contact is business-as-usual in the long run. A third scenario will consider slow decontamination and also take into account that individuals keep the habit of respecting barrier gestures and corresponds in the simulations to 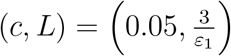. The contact is then a third of what it used to be before the epidemic outbreak. The three scenarios are plotted in Figure 3, using for disconfinement time *t* = 49 which corresponds to April the 15th.

**Figure 2:**
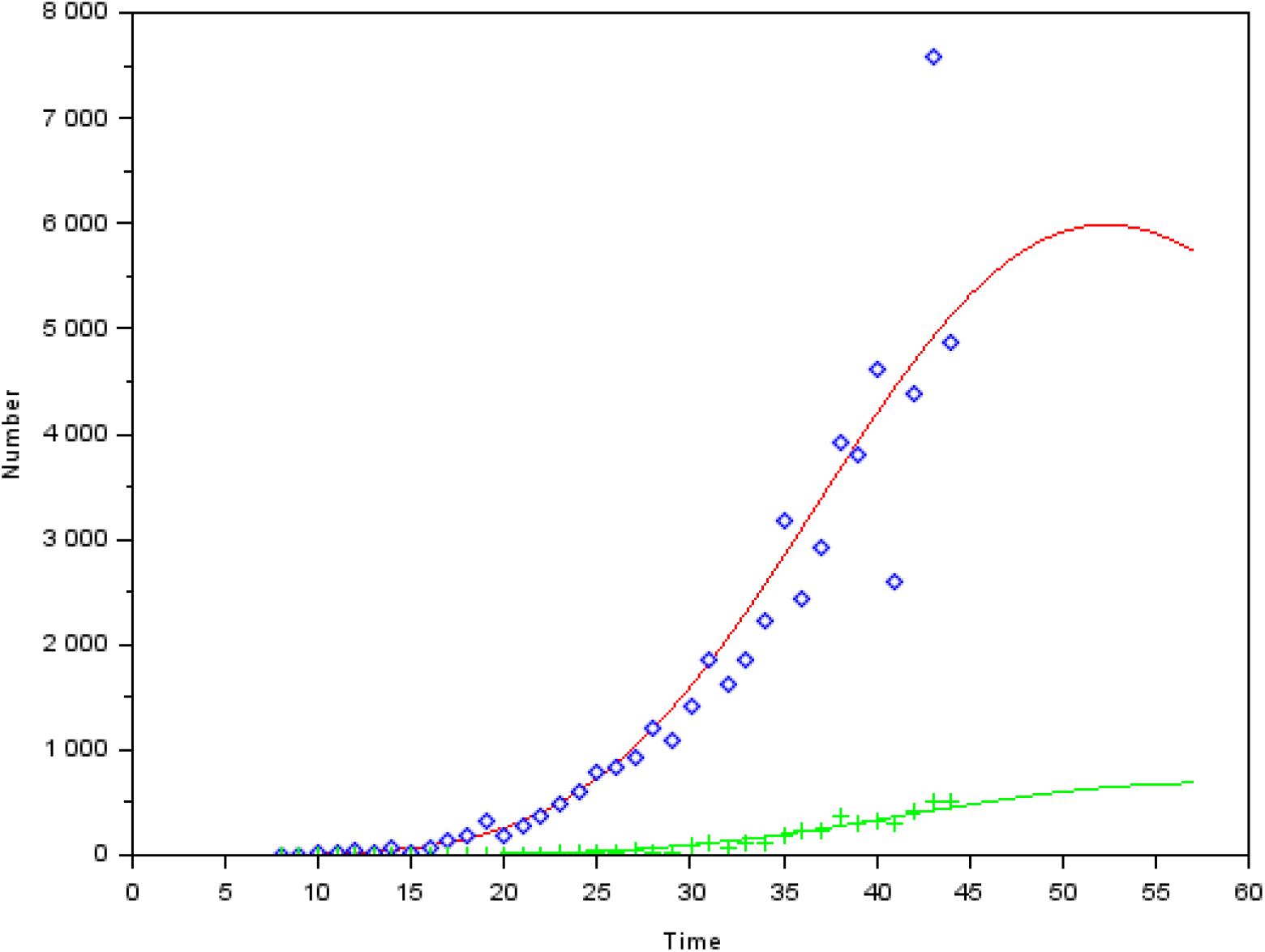
On this figure, we plot *t* → *v*_1_*E*(*t*) the number of daily new reported cases (red solid line) and data for this variable (blue dots). We also plot *t → γI*_*R*_(*t*) the daily number of new deaths and data for this variable (green crosses)

**Figure 3:**
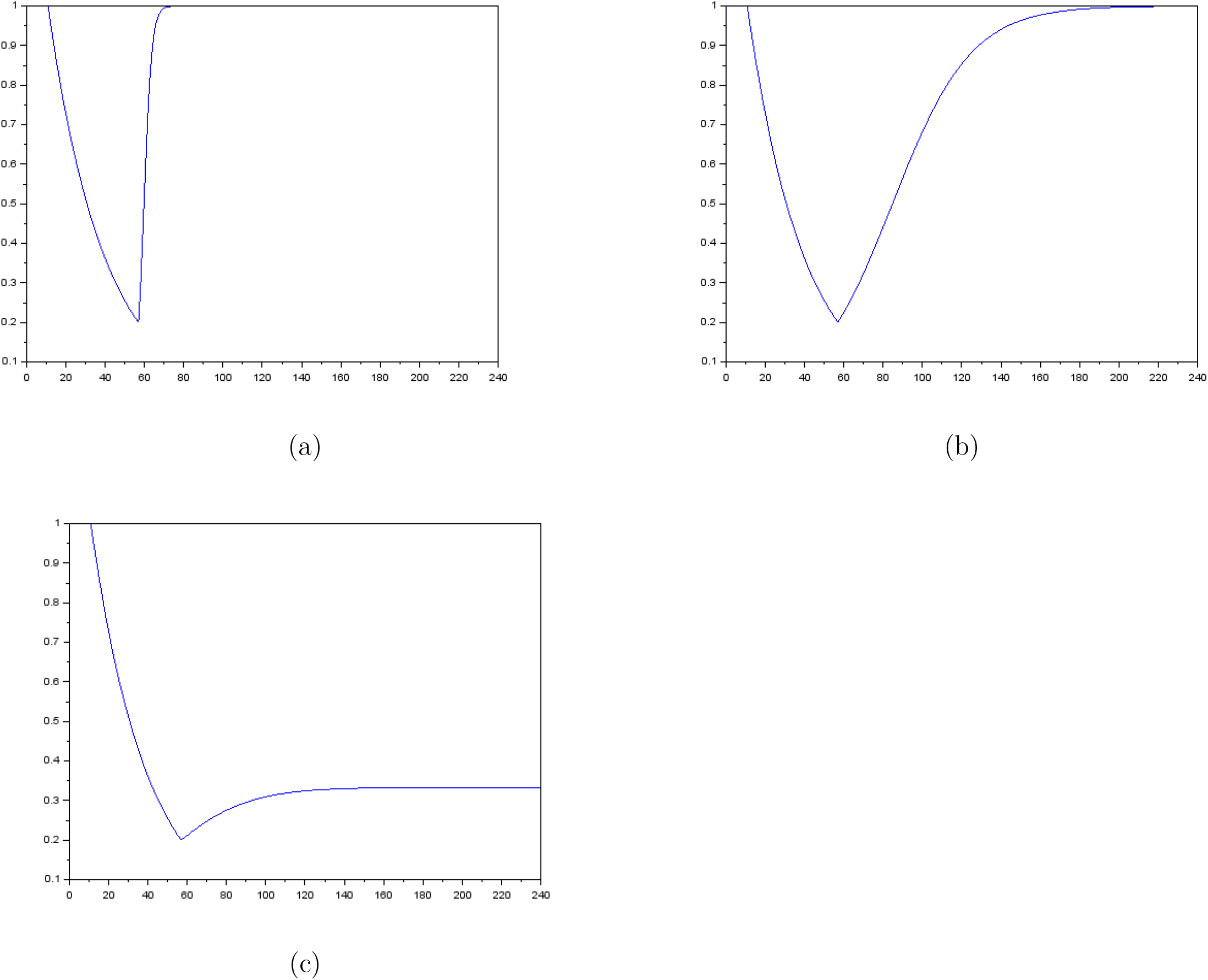
Fast disconfinement scenario and then business as usual (a). Slow disconfinement scenario and then business as usual (b). Slow disconfinement scenario and protection habit (c).

The trajectories of new reported cases and deaths are presented in Figure 4 under the various scenarios. It can be seen that with the business-as-usual scenario (with either fast or slow disconfinement) a major peak of the epidemics appears in spring (Figure (a) and (b)). The only difference between fast and slow disconfinement is the timing of the peak, which arises sooner in case of fast disconfinement and its magnitude, which is higher as fast disconfinement is concerned. Taking into account that individuals will reduce their contact habit from one third is not enough to prevent a major epidemic outbreak that would happen in November.

**Figure 4:**
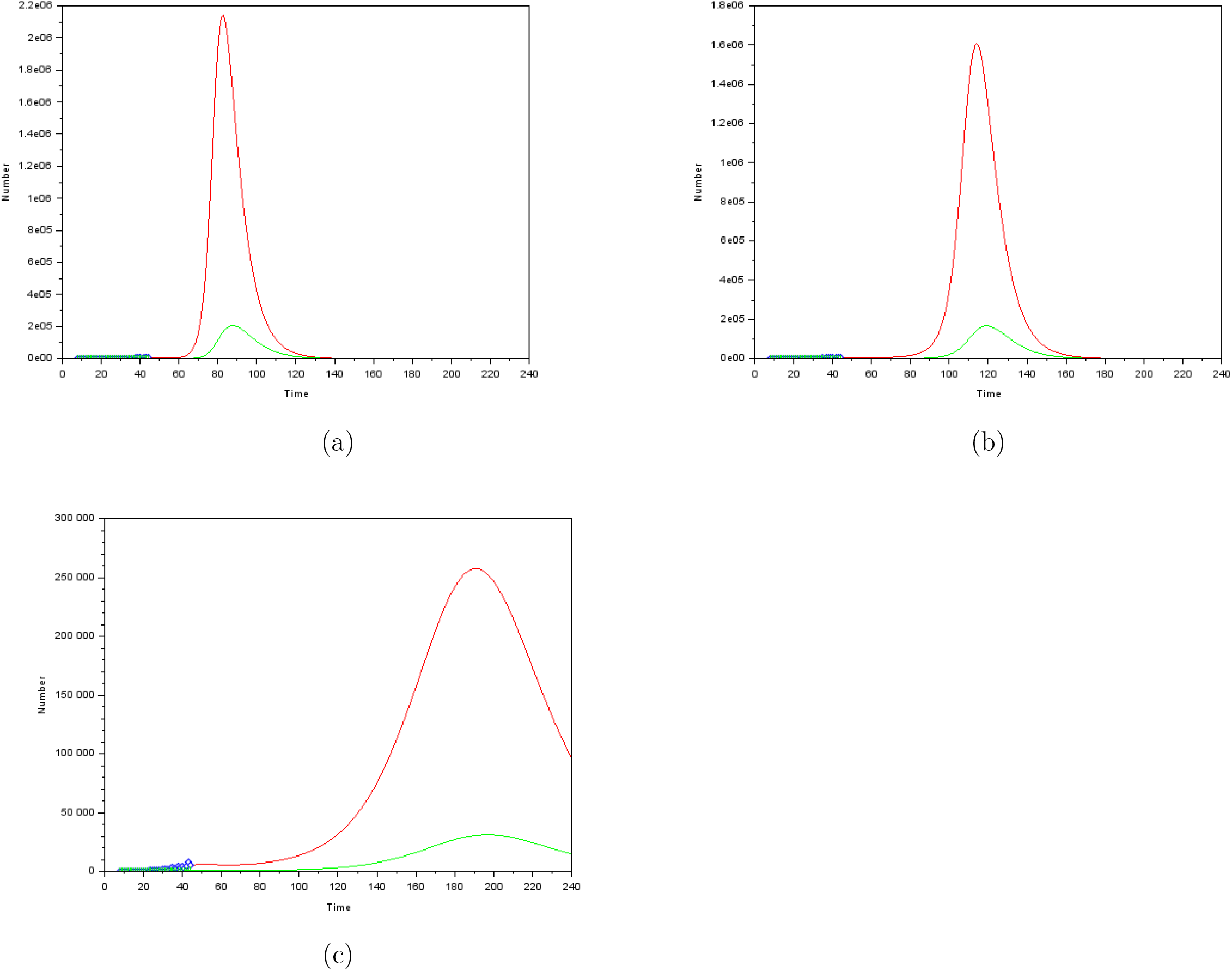
Fast disconfinement scenario and then business-as-usual (a). Slow disconfinement scenario and then business-as-usual (b). Slow disconfinement scenario and barrier gestures habit (c).

**Figure 5:**
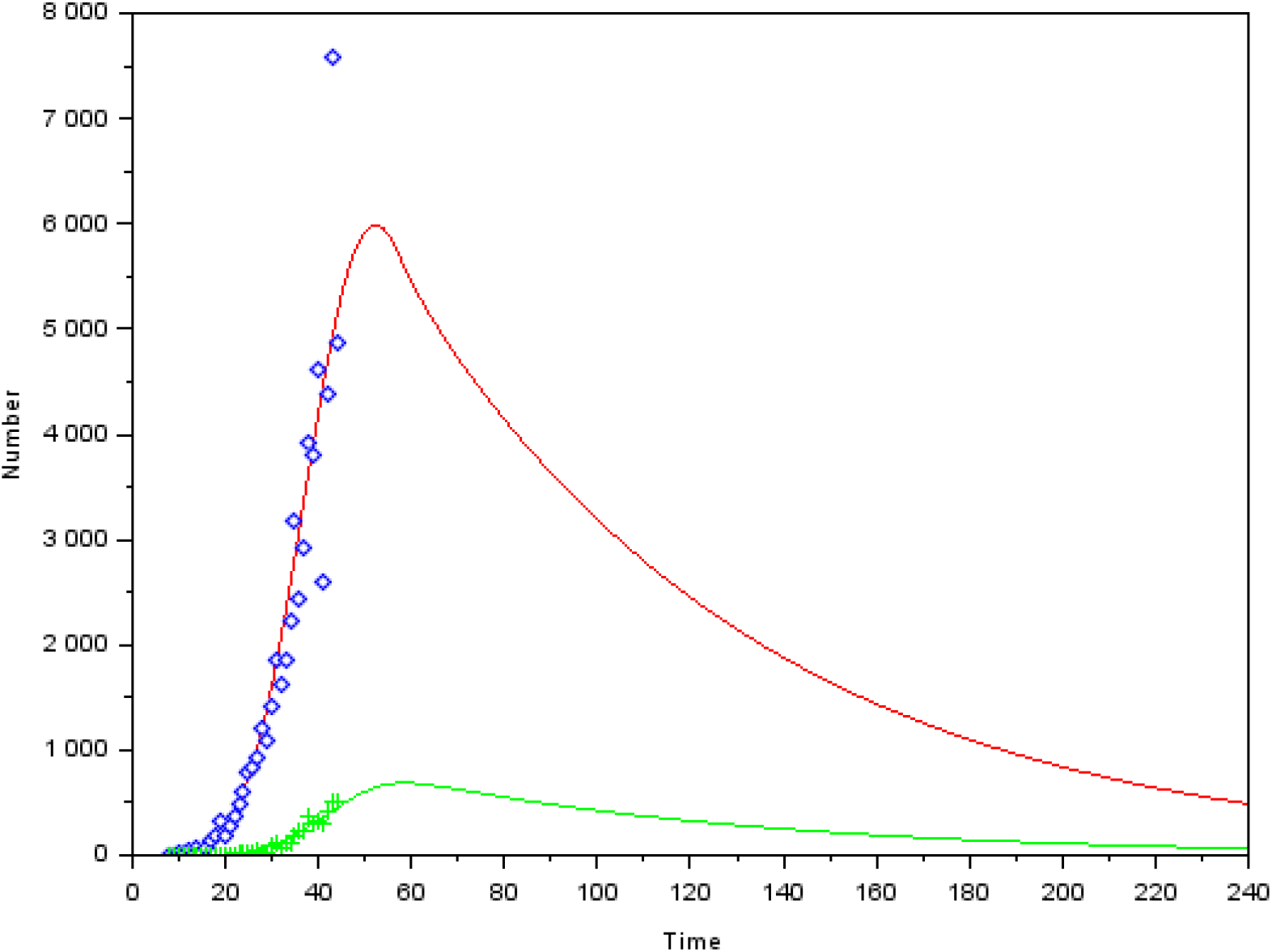
Slow disconfinement scenario and drastic reduction of contacts in the future.

If we consider a fourth scenario with slow disconfinement and reducing contact from a fifth, that corresponds in the modeling to 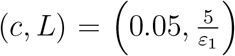, the re-emergence of the epidemics in autumn does not happen. Over this threshold, there is re-emergence of the epidemics. This threshold correspond to the reduction of contact which would remain the same than the one achieved by the 15th of April.

## 2 Discussion

France was the 7th country hit by the COVID-19. According of the experience of foreign governments who were already fighting the epidemics in their own country, it has been decided to establish strict confinement in the early stage of the epidemics. It has not prevented deaths and numerous new reported cases however but it has slow down drastically the course of the epidemics. The economic loss is huge (production has decreased from one third) and confinement is only a temporary measure. This is why disconfinement measures have been studied in this paper. We have seen that even strict disconfinement scenario does not prevent from a major epidemic outbreak either in spring, either in august.

The disconfinement process and the public policy communication to help individuals to keep barrier gestures habit may be improved by the use of a more intensive screening. In Figure 6, a more intense screening process is taken into account together with scenario 3. It can be seen that it would avoid the re-emergence of the disease.

An other health policy measure might be to delay the date of disconfinement. Three postponements of the end of confinement are proposed in Figure 7. A one week delay has few impacts on either the timing of the re-emergence or on its magnitude, as it can be seen on Figure 7(a). A one month postponement delays the re-emergence of one year, which is really significant but has few impact on the magnitude of the second outbreak. A two month delays has a major impact on both the timing and the magnitude of the epidemics

**Figure 6:**
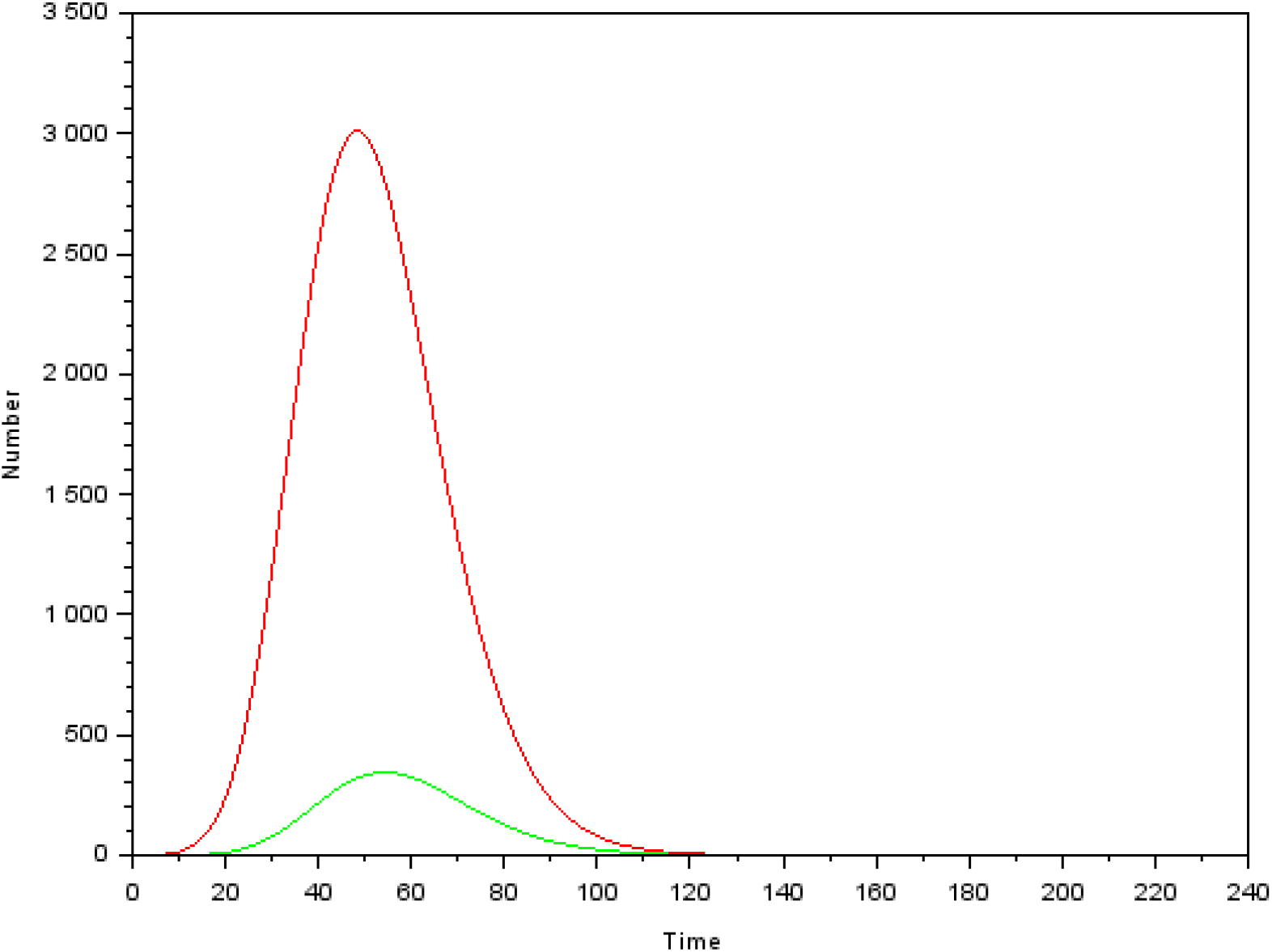
Slow disconfinement scenario and barrier gestures habit 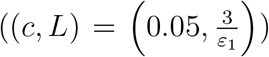 together with *f* = 0.8 (instead of *f* = 0.6 previously).

**Figure 7:**
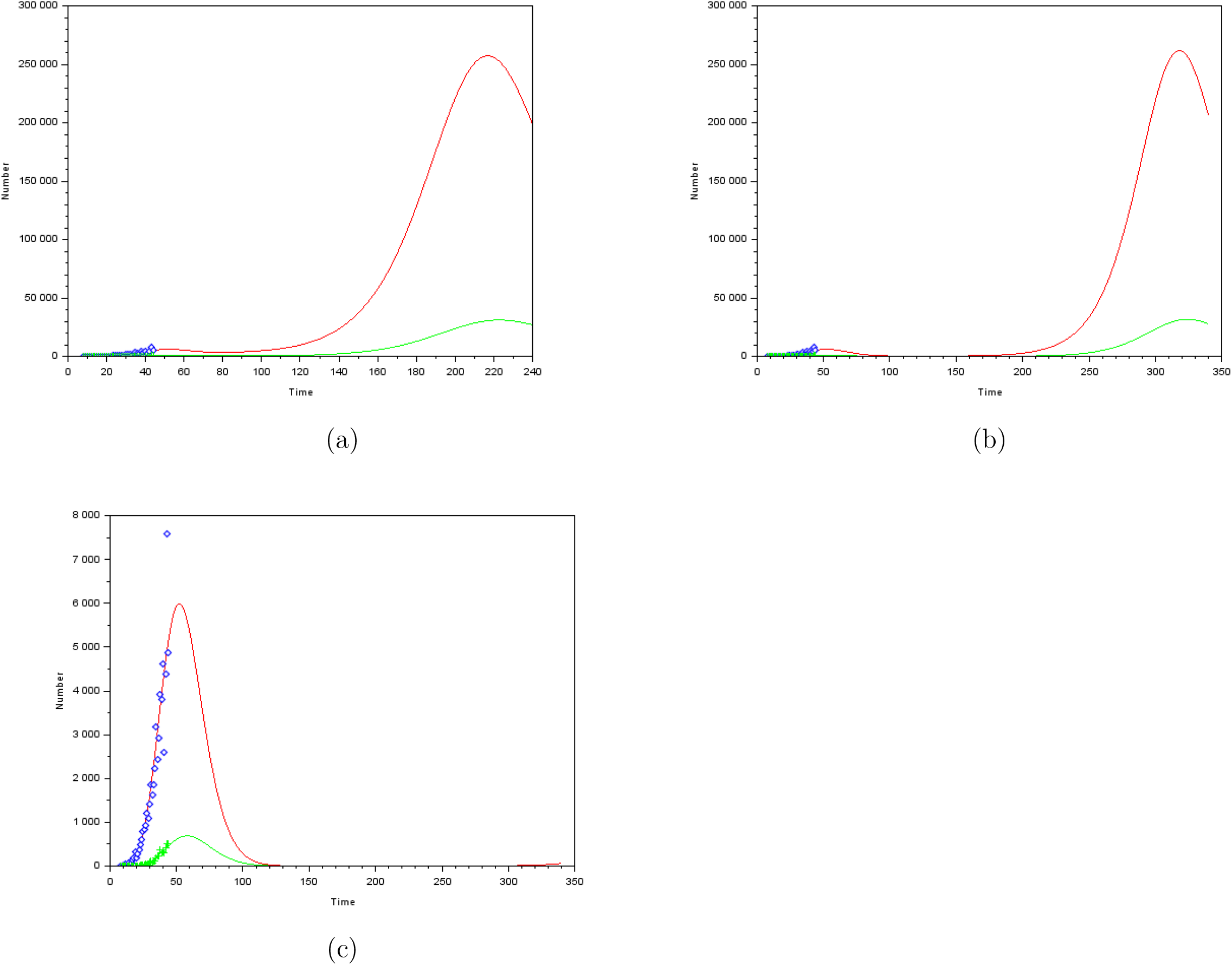
disconfinement on April 22nd (a). disconfinement in May the 15th (b). disconfinement in June the 15th (c).

## 3 Supplementary material

### 3.1 Determination of *Y*

We use the notations of Arino et al. [1]. Let us consider

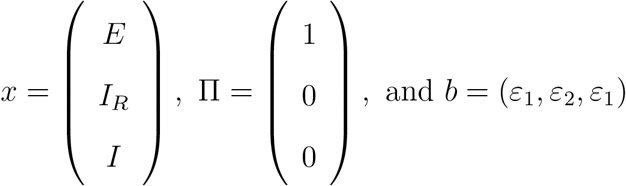

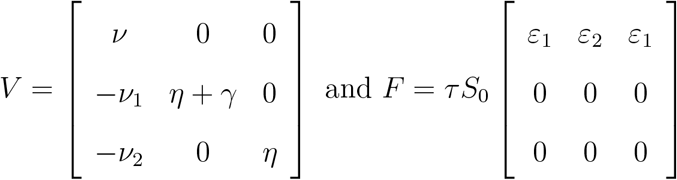

Since the model includes confinement policy *ε*_1_, we would call *ρ* (*FV* ^−1^) the *control* reproduction number and denote *ℛ*_*C*_ = *ρ* (*FV* ^−1^). *ℛ*_*C*_ can be explicitly computed as

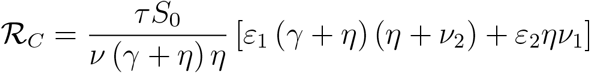

Let us consider *Y* a weighted number of infective which is defined as 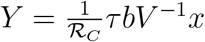. Thus *Y* is explicitly given by *Y* = *α*_1_*E* + *α*_2_*I*_*R*_ + *α*_3_*I*

Where

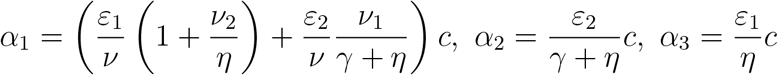

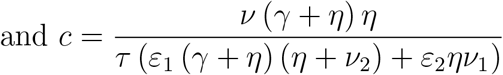

Then, taking the derivative of *Y* yields

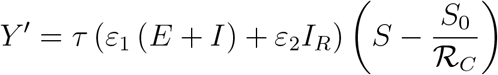

Thus

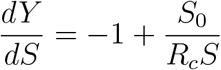

which has an explicit solution. Denoting *S*_∞_ = lim_*t*→∞_ *S* (*t*) and *Y*_∞_ = lim_*t*→∞_ *Y* (*t*), and as *Y*_∞_ = 0, the final size relation is

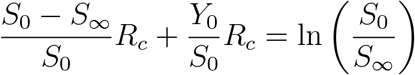

## Data Availability

I use public data from the French Public Agency of Health

## Footnotes

1 The computations are detailed in the Supplementary Material section

## Notes

### Competing Interest Statement

The authors have declared no competing interest.

### Funding Statement

No funding

